# Obstetrical safety indicators for preventing hospital harms: a scoping review protocol

**DOI:** 10.1101/19012153

**Authors:** A Conway, J Reszel, MC Walker, JM Grimshaw, SI Dunn

## Abstract

**Introduction:** Optimizing the safety of obstetric patient care is a primary concern for many hospitals. Identifying performance indicators that measure aspects of patient care processes related to preventable harms can present opportunities to improve health systems.

In this paper, we present our protocol for a scoping review to identify performance indicators for obstetric safety. We aim to identify a comprehensive list of obstetric safety indicators which may help reduce the number of preventable patient harms, to summarize the data and to synthesize the results.

**Methods and analysis:** We will use the methodological framework described by Arksey and O’ Malley and further expanded by Levac. We will search multiple electronic databases such as Medline, Embase, CINAHL and the Cochrane Library as well as websites from professional bodies and other organisations, using an iterative search strategy. We will include indicators that relate to preventable harms in the process of obstetric care.

Two reviewers will independently screen titles and abstracts of search results to determine eligibility for inclusion. For records where eligibility is not clear, the reviewers will screen the full text version. If reviewers’ decisions regarding eligibility differ, a third reviewer will review the full text record. Two reviewers will independently extract data from records that meet our inclusion criteria using a standardized data collection form. We will narratively describe quantitative data, such as the frequency with which indicators are identified, and conduct a thematic analysis of the qualitative data. We will compile a comprehensive list of patient safety indicators identified during our scoping review and organise them according to concepts that best suit the data such as the Donabedian model or the Hospital Harm Framework. We will discuss the implications of the indicators for future research, clinical practice and policy making.

We will report the conduct of the review using the Preferred Reporting Items for Systematic Reviews and Meta-Analyses extension for scoping reviews (PRISMA ScR) Checklist.

**Ethics and Dissemination:** The sources of information included in this scoping review will be available to the public. Therefore, ethical review for this research is not warranted. We will disseminate our research results using multiple modes of delivery such as a peer-reviewed publication, conference presentations and stakeholder communications.

**Strengths and limitations of this study**

1. We will use systematic methods to conduct and report this scoping review based on established methods introduced by Arksey and O’ Malley and further developed by Levac.
2. We will report the conduct of the review using guidance from the PRISMA Extension for Scoping Reviews (PRISMA-ScR).
3. We will limit the focus of this review to patient safety indicators from English language resources aimed at health system processes in developed countries.
4. We will not assess the risk of bias or certainty of the evidence in the included records because this is a scoping review to identify potential obstetrical safety indicators.
5. After the scoping review, we will present the identified indicators to key informants in a consensus process to create a shortlist that may not be suitable for inclusion in a future obstetrical safety dashboard for BORN (Better Outcomes Registry and Network) Ontario. BORN collects and safeguards data, and shares information to improve the delivery of maternal, neonatal and child healthcare across the province.

## Introduction

An aim of healthcare delivery is to avoid preventable harm to patients. However, human limitations and the complex nature of health care systems are contributing factors that make errors an expected phenomenon (1). A systematic review by Nabhan and colleagues found that the most frequently used definition of preventable harms is “presence of an identifiable, modifiable cause of harm” (2). Preventable harms that occur during hospital stays are also known as hospital harms, patient safety incidents, hospital adverse events or hospital errors. The Canadian Institute for Health Information (CIHI) defines hospital harms as the “rate of acute care hospitalizations with at least one occurrence of unintended harm during a hospital stay that could have been potentially prevented by implementing known evidence-informed practices” (3).

In 2000, the Institute of Medicine in the United States published a ground-breaking report *To Err is Human: Building a Safer Health System* (4) which raised awareness around errors in healthcare processes and their negative impact on patient morbidity and mortality. In 2004, the results of the Canadian Adverse Events Study showed that according to the sample of 3745 non-psychiatric, non-obstetric adult patient charts reviewed, the rate of preventable adverse events was 2.8% (95% confidence interval [CI], weighted, 2.0 – 3.6) and the mortality rate from preventable adverse events was 0.66% (95% CI, weighted, 0.37%–0.95%) (5). Since then, the Canadian Patient Safety Institute (CPSI) reported that patient harm is Canada’s third highest cause of death. Preventable harms occur in one out of every 18 hospital visits and resulting treatments costs CAD$2.75 billion annually (6). Regarding the field of obstetrics, the most recently published hospital harm results from the Canadian Institute for Health Information (CIHI) and the Canadian Patient Safety Institute (CPSI), (7) during the fiscal year 2017 – 2018, reported 5566 incidences of obstetric trauma caused by health care or medication-associated conditions and 4242 incidences caused by procedure-associated conditions. They also reported that during the same period, 670 incidences of obstetric hemorrhage were caused by health care or medication associated conditions and 960 incidences were caused by procedure associated-conditions. In addition, 855 birth trauma incidences were caused by health care or medication-associated conditions and 1417 incidences were caused by procedure-associated conditions.

Certain harms require exploration of each patient case to assess whether the harm is potentially preventable, and others are more obviously preventable, such as fatal medication doses (8) or wrong-site surgeries. The Hospital Harm Framework classifies harms into 4 categories: health care/medication-associated conditions; health care associated infections; patient accidents; and procedure-associated conditions (9).

Several studies have shown strategies that have successfully reduced preventable patient harm. Existing research has shown that making patient safety central to the delivery of care can lead to system-level improvements. In the United States, Pettker and colleagues implemented a comprehensive obstetric patient safety program which led to reductions in adverse outcomes and litigation cases, and improvements in staff perceptions of safety and improvements in the safety climate, (10–12). Also in the United States, the California Maternal Quality Care Collaborative [CMQCC] California has identified a correlation between the work of perinatal quality collaboratives (PQCs) and a 50% reduction in maternal mortality rates in the state of California (13).

Audit and feedback interventions may help to prevent patient harm. The Institute for Healthcare Improvement (IHI) Framework for Safe, Reliable and Effective Care identifies processes of improvement and measurement as a part of the health care learning system (14). In the UK, an ethnographic study of a maternity unit with an excellent record for patient safety found that staff partly attributed its success to a maternity dashboard which facilitated audit and feedback (15). The Better Outcomes Registry & Network (BORN Ontario) developed a dashboard of six key clinical performance indicators to measure data relating to the quality of maternal and newborn patient care in Ontario, Canada (16–18). In a pilot study, a quality improvement project focusing on one of the indicators identified by BORN, led to a welcome and significant reduction in the rate of elective repeat caesarean sections (ECRS) in low risk women at less than 39 weeks’ gestation (19). In an interrupted time series study that evaluated post-implementation effectiveness of the maternal newborn dashboard, the researchers found that it led to improvements in four of the six key indicators (20). A systematic review by Ivers and colleagues (21) found that audit and feedback can lead to small improvements in the practice of care delivery by health care professionals. In a rapid review, Anthony and colleagues found that audit and feedback in combination with other obstetrical safety initiatives may reduce maternal and neonatal morbidity and mortality (22).

It is necessary to identify patient safety indicators to ensure the effectiveness of future audit and feedback strategies. Hearnshaw and colleagues have highlighted the need for useful audit review criteria that are developed or selected using systematic methods (23,24). However, in the field of obstetrics, it is currently unclear which indicators relate to, or have the potential to improve obstetric safety. In this scoping review, we want to contribute to the improvement of obstetric patient safety by identifying indicators as the starting point of an approach to harm reduction.

The term indicator is generally understood to mean a measure that provides information on health service delivery across organisations which enables tracking and comparison of performance over time. In a paper first published in 1966, Donebedian identified three types of measures of quality which can be used to evaluate the process of care at the patient-healthcare provider level; outcome measures; process measures and structure measures (25). A fourth type, balancing measures, relate to positive or negative phenomenon which happen as an unintended consequence of a change (14).

Many desirable criteria for measuring the quality of care delivered by a health system have been proposed in the literature. Kessner and colleagues (26) suggested using tracers which are a set of specific health conditions or activities. They identified the following criteria for tracers: having the potential to be positively impacted by health care activities, clearly definable and easily diagnosable, have a level of prevalence that leads to data collection adequate for statistical analysis, having a demonstrable potential to improve health conditions, relate to processes involving prevention, diagnosis, treatment or rehabilitation, and contextual influences should be understood. Janakariaman and Ecker listed their ideal criteria for measures as: easily definable and observable; important to patients and health care professionals; identifies areas where improvements are needed and uses accessible data (27).

A systematic review by Saturno-Hernández and colleagues found that despite the existence of a large number of indicators for monitoring obstetric care, they were lacking in scientific rigor and many are not suitable for implementation (28). They used the following desirable criteria for indicators: presenting a full description, clearly based on explicit evidence, reliable, and feasible (confirmed by pilot testing).

Although it is likely that indicators have the potential to improve patient safety there are no consistent standards for measuring obstetric patient safety. When deciding on indicators it is extremely important to understand what you want to measure and how it can be measured effectively. In order to use obstetric safety indicators as part of a harm reduction strategy they should resonate with health care professionals.

Quality and safety indicators are closely linked and sometimes used interchangeably. However, safety indicators are a sub-type of quality indicators. For this review, we will use the definition of patient safety indicators used by Kristensen and colleagues i.e. measures that directly or indirectly monitor preventable hospital harms (29). For these indicators it is important to map indicators to goals which are actionable. In the field of obstetric patient safety, there is a need to identify indicators which measure the aspects of care that relate to preventable harms caused by medical errors. This can be challenging because it is not always clear which indicators should be used and there are variations in practice and outcomes across hospitals, regions etc.

It is important to note that some safety indicators may be judged as avoidable depending on the patient case. Therefore, we will focus on obstetric safety indicators that relate to the prevention of harms that are at least partly potentially avoidable to patients.

The aim of our review will be to scope the body of relevant literature (30) to identify a comprehensive list of potential obstetric safety indicators which can help reduce preventable patient harms. The objectives of the scoping review are to search for and identify records discussing indicators that can optimize obstetrical safety, to summarize the data and to synthesize the results. This scoping review is being undertaken to inform which indicators will be included in a patient safety dashboard from BORN Ontario.

## Methods and analysis

We will undertake a scoping review to systematically map the available literature on performance indicators relevant to the reduction of preventable harms within the purview of obstetric safety. We will use the PRISMA extension for scoping reviews PRISMA ScR Checklist (31) to guide the reporting of the scoping review. We will use the scoping study framework introduced by Arksey and O’ Malley (32) and the recommendations later developed by Levac (33) to guide our methods. We will follow all six suggested stages as described below.

### Stage 1: Identifying the research question

Through consultation with the author team we defined the concepts, target population and type of indicators of interest, and iteratively developed the main research question for the scoping review. We ensured that it aligned with the purpose and scope of the review and the expected outcome (a comprehensive list of relevant indicators).

This question is: *According to the available literature, what are the obstetrical safety indicators related to processes of care for low risk births that aim to reduce preventable hospital harms?*

For the purposes of this review, we will use the Southern Ontario Obstetrical Network (SOON) definition of a low-risk birth target population namely, nulliparous pregnant individuals, singleton gestation with cephalic presentation, who deliver at 37 weeks of gestation or more (34). SOON has also specified exclusion criteria shown in the table in Appendix 2. The term ‘fetal anomalies’ featured in the SOON definition is an umbrella term for multiple anomalies. Therefore, we will use the list of major sentinel congenital anomalies (see Appendix 3) that was developed in 2017 by BORN Ontario and OCAC (Ontario Congenital Anomalies Committee) as an additional tool during screening.

### Stage 2: Identifying relevant studies

The search strategy will be drafted by one of the authors who has trained as an information specialist (AC) and developed iteratively based on consultation with the author team and at least one other library and information professional. We will aim to be as comprehensive as possible while considering resource constraints (such as time) to avoid large volumes of search results. We will search electronic databases such as Medline (Ovid), Embase Classic + Embase 1947 to 2019 (Ovid), CINAHL (EBSCO), the Cochrane Library via www.cochranelibrary.com. We will search for grey literature resources such as those identified in the Canadian Agency for Drugs and Technologies in Health (CADTH) grey literature checklist *Grey Matters: a practical tool for searching health-related grey literature* (35). We will also search relevant websites of professional bodies or organisations such as the Society of Obstetricians and Gynaecologists of Canada (SOGC)(36), Royal College of Obstetricians and Gynaecologists (RCOG)(37), Royal Australian and New Zealand College of Obstetricians and Gynaecologists (RANZCOG) (38) and American College of Obstetricians and Gynecologists (ACOG) (39) etc. We will contact individuals or groups to source additional material if deemed appropriate.

We will use both thesauri terms (such as MeSH, Emtree and CINAHL headings) and free text terms as well as Boolean operators and field limiters (where appropriate), to build our search. We will report our conduct of the search including information such as: each database or source of information; dates of the searches; applied limits or filters; the number of records found (for each database or source of information) and search terms from at least one electronic database. We will undertake reference searching and citation searching for records which meet our inclusion criteria.

See Appendix 1 for our currently proposed search strategy for Medline which may be further developed before it is implemented. It will be tailored according to the functionality of each database or other resource we search.

### Stage 3: Selecting studies

Two reviewers will undertake a test of interrater reliability to increase the likelihood of reliable screening. We will select 50 records and screen them for inclusion. We will calculate Cohen’s kappa (40). We may revise the inclusion and exclusion criteria if indicated. When we have achieved a 0.80 or higher level of agreement between raters, we will independently conduct three phases of screening. Firstly, we will screen the titles and abstracts of records that are not randomized trials to determine eligibility for inclusion against the agreed inclusion and exclusion criteria using screening software (Covidence). Secondly, we will screen 30 randomised trials to see if they identify additional indicators. If this step is not useful, we will not screen the remaining randomised trials. Thirdly, two reviewers will independently screen the full-text records for which it was not clear whether they met inclusion criteria in the previous screening phase. If there is disagreement regarding the inclusion of a record(s), another reviewer will assess the full text. We will undertake interrater reliability testing after every 1,000 records screened.

Records will be selected using the following inclusion criteria:

- one or more indicator is proposed or in use to prevent harm and improve obstetric safety
- specifies or includes a low risk birth population
- in-hospital settings
- care during the period from admission to discharge from hospital following birth
- indicators that can be improved or are controllable
- feature a level of measurability/auditability

We will include any type of primary study and most other record types discussing indicators such as reports, guidance documents and discussion papers, with the exception of conference papers. We will not exclude sources based on the year they were made available or published, or on their publication status (published or unpublished). We will only include English language records due to resource restrictions for translation services. We will exclude records focusing on indicators specifically designed for use within underdeveloped or developing world settings because our indicators should be applicable to a developed country (Canada).

We will present an adapted PRISMA flow diagram (41) to document the number of records identified, screened, assessed for eligibility, and included or excluded. We will document the main reasons why records were excluded.

### Stage 4: Charting the data

We will identify variables that are relevant and develop a standardized data collection form (15). Two reviewers will independently carry out data extraction for each included record. Initially, we will independently extract data from five randomly-selected, included records and compare the extracted data for consistency. The form may be iteratively developed throughout the charting process. Data items will include:

- indicator source (bibliographic information)
- study design and/or record format
- country of origin
- type of indicator (outcome/process/structure/balancing)
- indicator name
- indicator definition
- numerator
- denominator
- measurement properties (tools, instruments etc.)
- the population that is targeted by the indicator; maternal, fetal or infant following birth (or a combination)
- the timeframe covered by the indicator; admission for birth including care during labour, during birth, during the postpartum period prior to discharge from hospital
- category of hospital harm being addressed
- whether indicator relates to a desirable or undesirable event
- evidence-based indicator (yes/no/unclear)

### Stage 5: Collating, summarizing and reporting results

As recommended by Levac, we will divide this final stage into three parts: analysis of data; reporting of results and discussion of the implications of the results for research, practice and policy (33).

1. We will produce descriptive numerical summaries through analysis of the quantitative data, such as the frequency with which indicators are identified. The qualitative data will be uploaded to NVivo software and thematic analysis will be conducted.
2. We will report our results by compiling a comprehensive list of patient safety indicators identified during our scoping review and organizing them according to concepts that best suit the data such as the Donabedian model or categories within the Hospital Harm Framework.
3. We will further explore the implications of the indicators for further research, on clinical practice within obstetric units and how policy analysts or policy-makers may find the results useful.

### Stage 6: Consultation

This stage is presented as optional by Arksey and O’ Malley (32) and Levac and colleagues argue that it should be considered a necessary element of the methodological framework. Data from our scoping review will be fed into a future consensus process. This scoping review is the first in a multi-step approach to inform the development of a core set of indicators. It is unclear which measures will be considered important to stakeholders so the results of the research will be disseminated to a panel of experts in a consensus process. They will prioritize indicators and we will produce a shortlist with the potential to inform the development of a future obstetrical safety dashboard in the Ontarian health system. As an electronic audit and feedback system, the dashboard could be used to capture, monitor and report surveillance data entered by health care professionals, and aim to contribute to improvements in obstetric safety.

## Data Availability

The data will be made publicly available on the Open Science Framework or by request at

https://osf.io

## Ethics and Dissemination

This scoping review will not require ethical approval because included sources are or can be made available to the public (42).

This research will be disseminated with the intention of reaching a wide audience through the use of multiple modes of delivery such as stakeholder engagement, publication(s) in a peer-reviewed journal and conference paper(s).

## Authors’ contributions

In consultation with all authors, AC wrote the first draft which SID and JR reviewed. JMG and MCW reviewed a second draft. All authors contributed to the planning and writing of this protocol and approved submission of the final manuscript to the publishing journal.

**Appendix 1:**
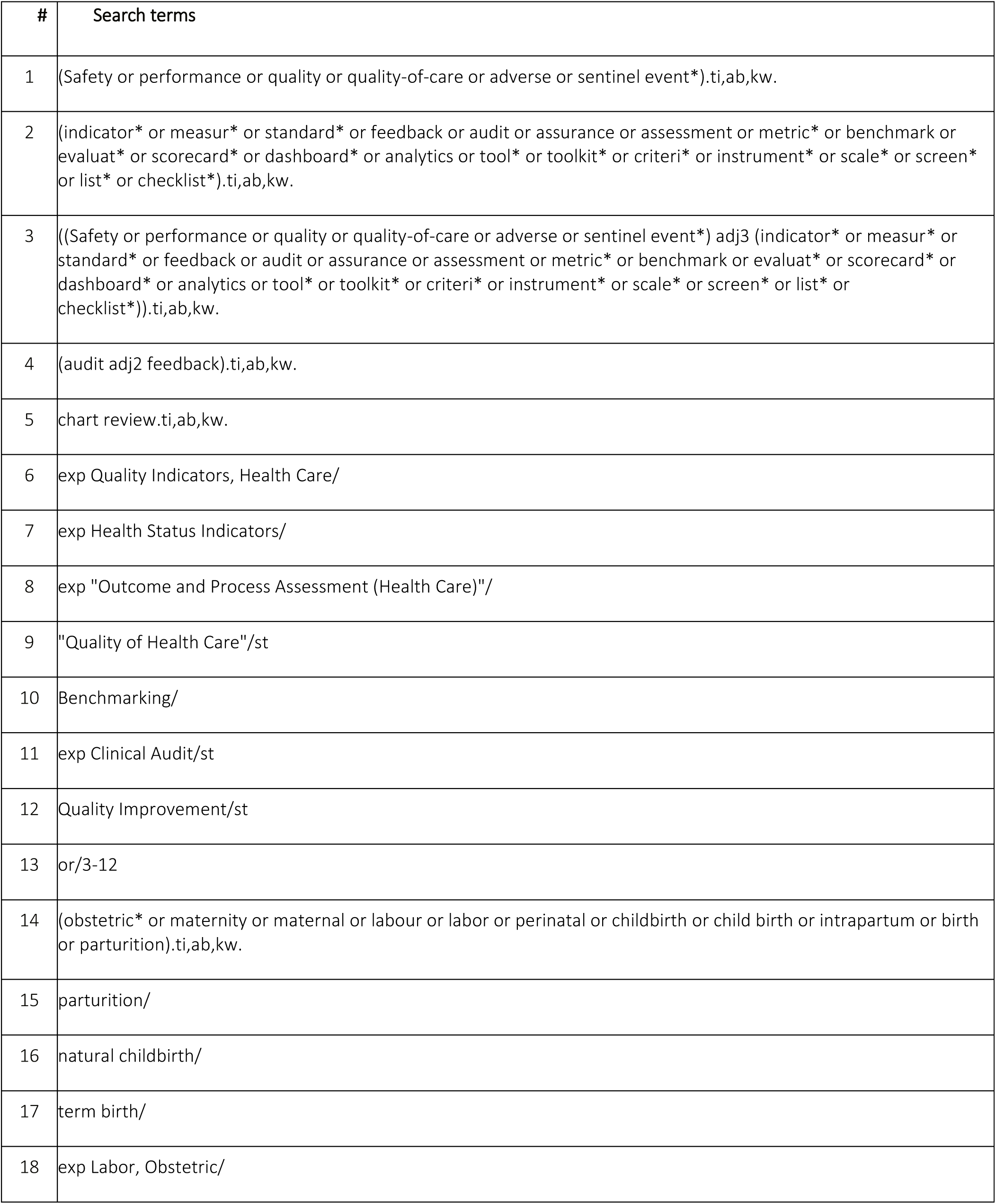
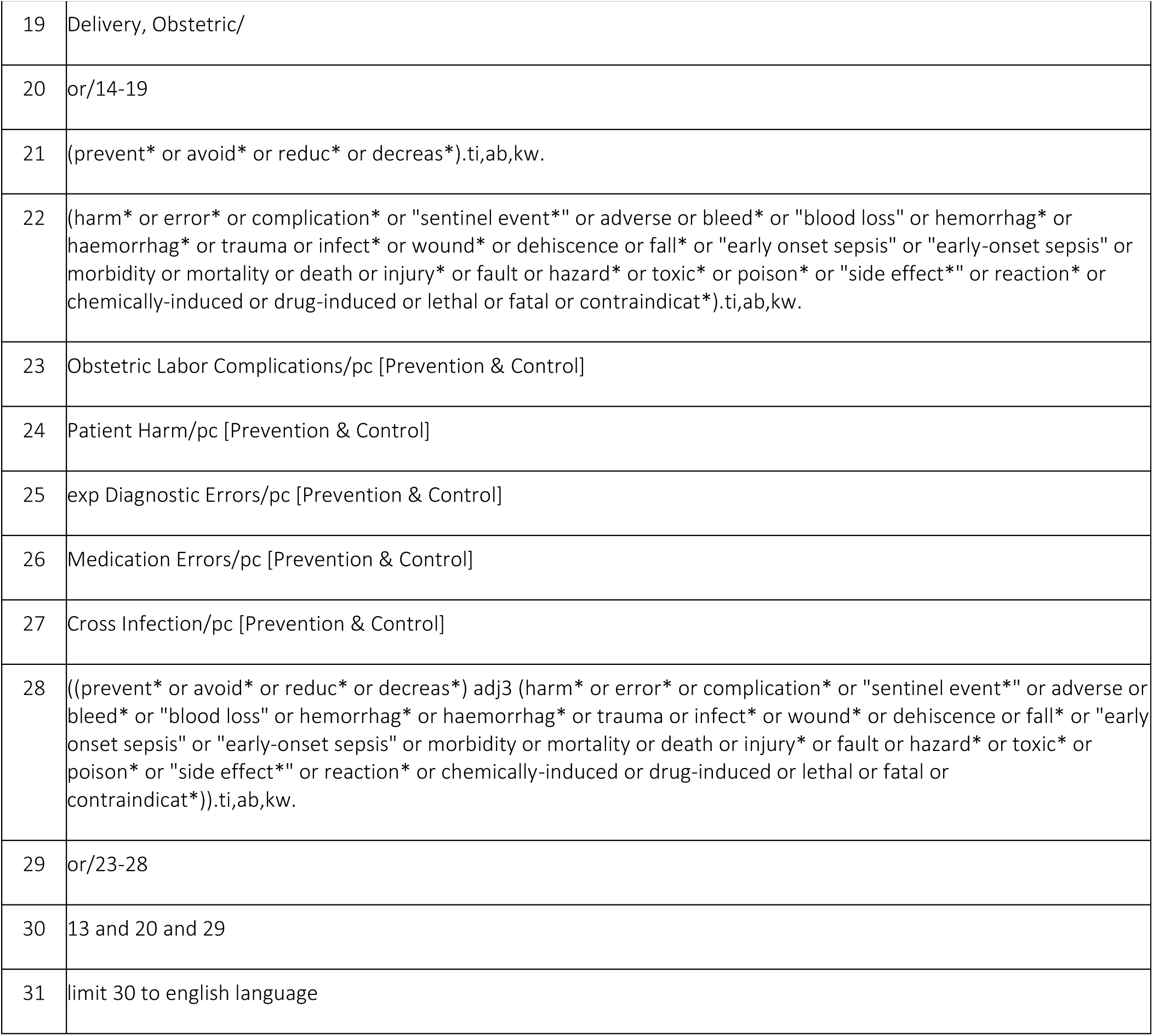
Search strategy for Medline (Ovid)

| # | Search terms |
| --- | --- |
| 1 | (Safety or performance or quality or quality-of-care or adverse or sentinel event*).ti,ab,kw. |
| 2 | (indicator* or measur* or standard* or feedback or audit or assurance or assessment or metric* or benchmark or evaluat* or scorecard* or dashboard* or analytics or tool* or toolkit* or criteri* or instrument* or scale* or screen* or list* or checklist*).ti,ab,kw. |
| 3 | ((Safety or performance or quality or quality-of-care or adverse or sentinel event*) adj3 (indicator* or measur* or standard* or feedback or audit or assurance or assessment or metric* or benchmark or evaluat* or scorecard* or dashboard* or analytics or tool* or toolkit* or criteri* or instrument* or scale* or screen* or list* or checklist*)).ti,ab,kw. |
| 4 | (audit adj2 feedback).ti,ab,kw. |
| 5 | chart review.ti,ab,kw. |
| 6 | exp Quality Indicators, Health Care/ |
| 7 | exp Health Status Indicators/ |
| 8 | exp "Outcome and Process Assessment (Health Care)"/ |
| 9 | "Quality of Health Care"/st |
| 10 | Benchmarking/ |
| 11 | exp Clinical Audit/st |
| 12 | Quality Improvement/st |
| 13 | or/3-12 |
| 14 | (obstetric* or maternity or maternal or labour or labor or perinatal or childbirth or child birth or intrapartum or birth or parturition).ti,ab,kw. |
| 15 | parturition/ |
| 16 | natural childbirth/ |
| 17 | term birth/ |
| 18 | exp Labor, Obstetric/ |
| 19 | Delivery, Obstetric/ |
| 20 | or/14-19 |
| 21 | (prevent* or avoid* or reduc* or decreas*).ti,ab,kw. |
| 22 | (harm* or error* or complication* or "sentinel event*" or adverse or bleed* or "blood loss" or hemorrhag* or haemorrhag* or trauma or infect* or wound* or dehiscence or fall* or "early onset sepsis" or "early-onset sepsis" or morbidity or mortality or death or injury* or fault or hazard* or toxic* or poison* or "side effect*" or reaction* or chemically-induced or drug-induced or lethal or fatal or contraindicat*).ti,ab,kw. |
| 23 | Obstetric Labor Complications/pc [Prevention & Control] |
| 24 | Patient Harm/pc [Prevention & Control] |
| 25 | exp Diagnostic Errors/pc [Prevention & Control] |
| 26 | Medication Errors/pc [Prevention & Control] |
| 27 | Cross Infection/pc [Prevention & Control] |
| 28 | ((prevent* or avoid* or reduc* or decreas*) adj3 (harm* or error* or complication* or "sentinel event*" or adverse or bleed* or "blood loss" or hemorrhag* or haemorrhag* or trauma or infect* or wound* or dehiscence or fall* or "early onset sepsis" or "early-onset sepsis" or morbidity or mortality or death or injury* or fault or hazard* or toxic* or poison* or "side effect*" or reaction* or chemically-induced or drug-induced or lethal or fatal or contraindicat*)).ti,ab,kw. |
| 29 | or/23-28 |
| 30 | 13 and 20 and 29 |
| 31 | limit 30 to english language |

**Appendix 2:**
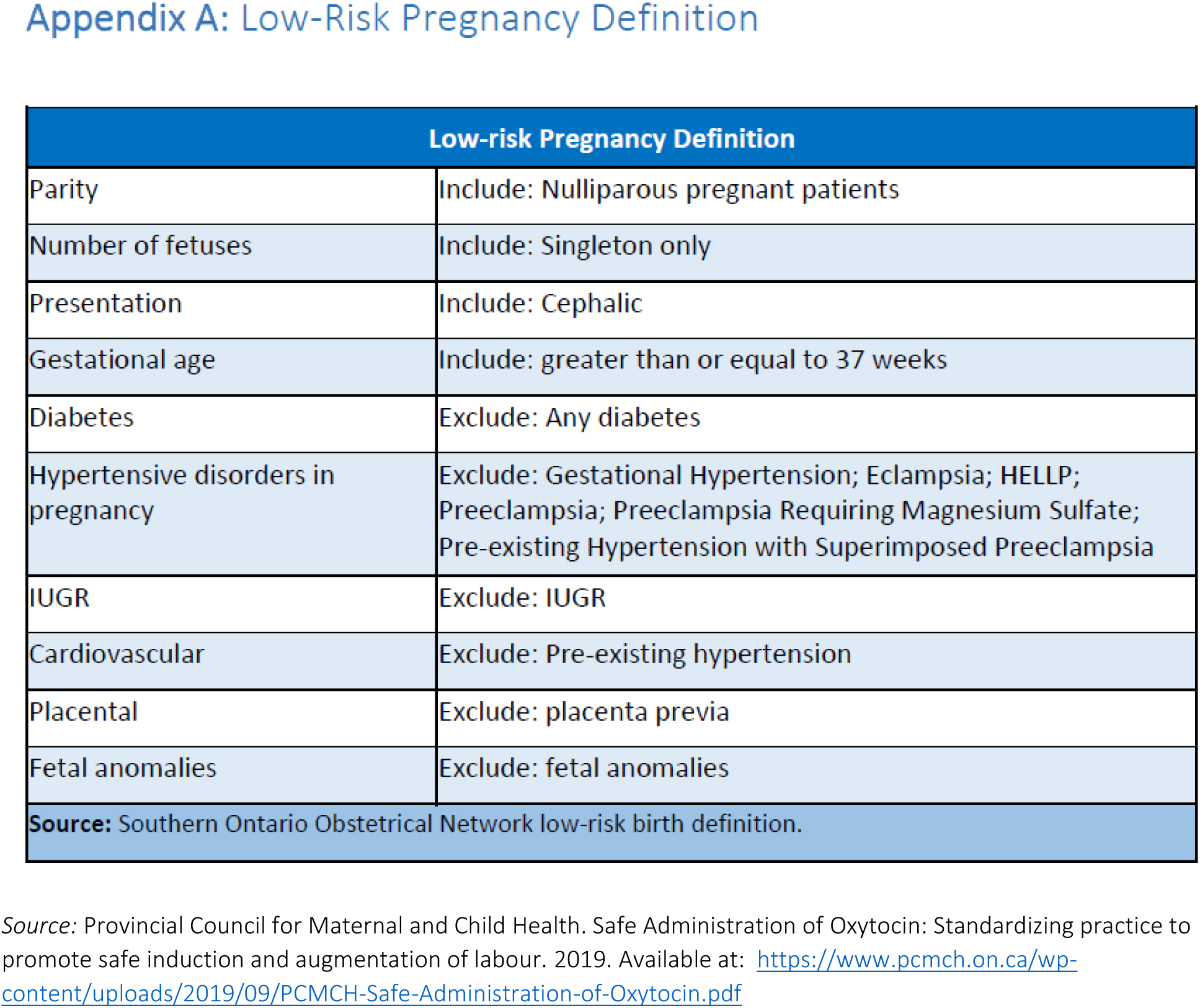
Southern Ontario Obstetrical Network (SOON) low-risk birth definition.

**Appendix 3:** Major sentinel congenital anomalies list

| System | Congenital Anomaly |
| --- | --- |
| Neural | 1. Anencephaly |
|  | 2. Acrania |
|  | 3. Craniorachischisis |
|  | 4. Encephalocele |
|  | 5. Hydranencephaly |
|  | 6. NTD with hydrocephalus |
|  | 7. NTD without hydrocephalus |
|  | 8. Hydrocephalus |
|  | 9. Atresia of foramina of Luschka and Magendie |
|  | 10. Dandy-Walker malformation / variant (DWM) |
|  | 11. Dandy-walker syndrome |
|  | 12. Aqueductal stenosis |
|  | 13. Hydrocephalus X-Linked |
| Cleft Lip / Palate | 14. Cleft lip & palate |
|  | 15. Cleft lip |
| Tracheo-Esophageal | 16. Esophageal atresia |
|  | 17. Tracheoesophageal fistula (TEF) |
| Small & Large Intestinal | 18. Atresia small or large intestine (NOTE: *\*cannot separate small & large intestinal atresia in BIS*). |
|  | 19. Duodenal atresia |
|  | 20. Abnormal small or large bowel |
|  | 21. Double bubble |
|  | 22. Bowel obstruction small or large intestine |
|  | 23. Stenosis small or large intestine |
|  | 24. Imperforate anus |
| **Hypospadias** | 25. Hypospadias (NOTE: * BIS does not collect epispadias). |
| **Limb Reductions** | 26. Limb reduction defect(s) (LRD) - lower limb |
|  | 27. Limb reduction defect(s) (LRD) - upper limb |
|  | 28. Radial ray anomaly (absent thumb) |
|  | 29. Adactyly (absent fingers/ toes) |
|  | 30. Ectrodactyly (lobster-claw / cleft hand) |
| **Gastroschisis** | 31. Gastroschisis |
| **Omphalocele** | 32. Omphalocele |
| **Renal Agenesis** | 33. Renal agenesis |
| **Heart** | 34. Hypoplastic left heart syndrome (HLHS) |
|  | 35. Tetralogy of Fallot (TOF) |
|  | 36. Transposition of great vessels (TGA) |
|  | 37. Transposition of great arteries - congenitally corrected (CCTGA) |
|  | 38. Hypoplastic left heart syndrome (HLHS) |
| **Trisomies** | 39. Trisomy 21 (Down syndrome) |
|  | 40. Trisomy 21 (Down syndrome) - translocation |
|  | 41. Trisomy 21 (Down syndrome) - mosaic |
|  | 42. Trisomy 18 |
|  | 43. Trisomy 13 |

Source: list jointly created by BORN Ontario and the OCAC (Ontario Congenital Anomalies Committee) in 2017.

